# A cross-sectional study of psychosocial factors and sickness presenteeism in Japanese workers during the COVID-19 pandemic

**DOI:** 10.1101/2021.07.23.21260909

**Authors:** Masashi Masuda, Tomohiro Ishimaru, Ayako Hino, Hajime Ando, Seiichiro Tateishi, Tomohisa Nagata, Mayumi Tsuji, Shinya Matsuda, Yoshihisa Fujino, for the CORoNaWork Project

## Abstract

**Background:** We examined the <u>association</u> between socioeconomic and health status, and lifestyle and sickness presenteeism among Japanese workers during the COVID-19 epidemic.

**Methods:** A cross-sectional study using an <u>Internet-monitor survey</u> was conducted in December, 2020 in Japan. Of 33,302 survey participants, we analyzed 27,036 <u>participants</u> (13,814 <u>men</u> and 13,222 <u>women</u>) who reported experience with sickness presenteeism.

**Results:** The <u>odds ratio (</u>OR) of sickness presenteeism associated with unmarried versus married status was 1.15. Respective figures for other variables were 1.11 for manual laboring work compared to desk work; 1.79 and 2.29 for loss of employment at the time the pandemic began and continuation of unemployment compared to maintaining employment during the pandemic; and 3.34 for a feeling of financial instability compared to stability.

**Conclusion:** The issue of sickness presenteeism has <u>become more prominent under the</u> COVID-19 <u>epidemic</u>.

## Introduction

Sickness presenteeism has rapidly attracted attention in occupational health. It has been defined as “the phenomenon of people, despite complaints and ill health that should prompt rest and absence from work, still turning up at their jobs”^1^. <u>Sickness presenteeism</u> has been reported in a number of countries^2-6^. It is known that workers with sickness presenteeism are aware of suboptimal general health and at increased risk of developing coronary artery disease and depression^7-9^. Known workplace impacts of sickness presenteeism include increased long-term leave, reduced work capacity, and the spread of infectious diseases^10-12^.

Vulnerable socioeconomic conditions, insecure employment status, as well as individual health views, work attitudes, company leave systems and culture influence workers’ experience of sickness presenteeism^13^. Workers who are worried about their job security are reluctant to take time off work for treatment. Workers with insecure employment are more likely to engage in sickness presenteeism because they fear that complaining about their health condition will be detrimental to the status of their employment contract. The shortage of employees and inadequate leave system also lead to an increase in sickness presenteeism^14,15^. Sick leave rates are low in areas where unemployment rate is high^16^, and it is thought that the fear of losing employment and poverty have led to situations in which people feel they have no choice but to work even if they are in poor physical condition^17^. In addition, worker behaviors, such as a positive job attitude, feelings of strong obligation, and considering absence a less legitimate option, are also known to result in sickness presenteeism^18^.

We hypothesized that the <u>coronavirus disease 2019 (</u>COVID-19) epidemic would affect worker experience of sickness presenteeism via an effect on their socioeconomic status and employment instability, as well as on their health behaviors. Shortly after the initial confirmation of COVID-19 in China, the infection spread globally, and its presence in Japan was confirmed in January 2020. <u>Since then, COVID-19 has continued to have a profound impact on healthcare systems, economic activities and people’s lives around the world. Under such circumstances, the impact of the COVID-19 epidemic on sickness presenteeism is considered to be an occupational hygiene issue.</u>

During a COVID-19 epidemic, workers may be more hesitant to report their health condition to the company when they are not feeling well. Reports worldwide have also noted the interruption of treatment for previously controlled diseases during the COVID-19 outbreak^19,20^, which also acts to increase sickness presenteeism. Furthermore, the management of sickness presenteeism is important in terms of preventing the spread of infection. Among various measures taken to halt the spread of COVID-19 in the workplace, self-reporting of physical condition and restriction of attendance by employees who have fever or other health problems are particularly important - the fact that some employees still come to work despite feeling ill is now an issue.

Some studies have considered the assumption that the COVID-19 epidemic affects the experience of workers’ sickness presenteeism^21,22^. <u>An analysis that combined survey data from 20,974 employees collected in Europe with Eurostat’s regional unemployment data shows that high unemployment rates enhance the presenteeism of disadvantaged workers^21^. The COVID-19 pandemic and consequent increase in labor market instability can increase the behavior of presenteeism. An Internet survey conducted in Belgium and the Netherlands after 8 weeks of coronavirus lockdown showed that the respondents experienced considerable levels of stress and concern about their financial situation, and also cancelled/delayed general healthcare and productivity loss during the COVID-19 pandemic^22^. The study also estimated presenteeism during the 8 weeks, but did not show an association between presenteeism and the factors experienced by respondents.</u> However, there are few reports on the characteristics of workers experiencing sickness presenteeism during the COVID-19 epidemic. We examined the <u>association</u> between socioeconomic and health status, and lifestyle and sickness presenteeism among Japanese workers during the COVID-19 epidemic.

## Methods

### Study Design and <u>Participants</u>

A cross-sectional study was conducted from December 22 to 26, 2020 at the time of the third wave of COVID-19 infection in Japan. The study was conducted as an Internet-monitor survey. Details of the study protocol have been described elsewhere^23^. Briefly, the data were obtained from workers currently under an employment contract at the time of the survey, and categorized by prefecture, type of job, and sex. Of 33,302 survey participants, <u>215 respondents were excluded because they were deemed to have provided fraudulent responses by the surveying company (Cross Marketing Inc.,). Subsequently, 6,051 respondents determined to have provided invalid or erroneous responses were excluded (See reference 23 for details). Finally,</u> 27,036 people were included in the study. The present analysis included all participants who responded that they had attended work despite needing to recuperate at home.

This study was approved by the ethics committee of the University of <u>Occupational and Environmental Health, Japan (R2-079. February 5, 2021)</u>. Informed consent was obtained via a form on the survey website.

### Assessment of sickness presenteeism

<u>Sickness presenteeism</u> was determined using the following single-item question: “How many days have you worked (including at home) in the last 30 days under conditions in which you would really like to take a day off?” <u>To evaluate the sickness presenteeism during the third wave of COVID-19 in Japan, we set the target period to 30 days.</u> Respondents were asked to respond with <u>a specific</u> number of days<u>; based on the distribution of days with sickness presenteeism, we set the cut-off to 3 days.</u>

### Survey of <u>participants</u> socioeconomic conditions, health status, and lifestyle factors

The questionnaire was conducted via the Internet and enquired about the <u>participant’s</u> socioeconomic conditions. Specifically, the survey enquired about age, sex, marital status (married; unmarried; widowed/divorced), occupation (job mainly involves desk work; interpersonal communication; labor), educational background <u>(Junior high school; High school; Vocational school/College; University; Graduate school)</u>, <u>equivalised</u> income (household income divided by the square root of household size), job change or unemployment after April 2020 (no resignation or job change; transfer to another company; resignation and immediate start at a new job; unemployment for a period, but presently employed; retired and started a business), and perception of financial situation (very difficult; somewhat difficult; slightly difficult; comfortable).

With regard to health status and psychological factors, we asked about self-rated health, psychological distress, loneliness, presence of supportive friends, and having a health condition that requires company consideration to allow work. Psychological distress was addressed using the Kessler <u>Psychological Distress Scale</u> (K6)^24^, whose validity in Japanese has been confirmed^25^. <u>Participants</u> with a K6 score of 5 or higher were considered to have mild psychological distress. Loneliness was examined with the question: “Have you ever felt alone <u>in the past month?</u>”, with response options of: “never,” “a little,” “sometimes,” “usually,” and “always.” <u>This method follows previous studies that assessed loneliness with a single question</u>^26^. The presence of a health condition that required the employer’s consideration to permit work was surveyed using the question: “Do you require consideration or support from your company to continue working in your current health condition?”, with the three response options of: “no,” “yes, but I have not received support;” and “yes, and I have received support.”

The questionnaire also contained a variety of questions on lifestyle and work-related factors, including smoking (never; quit more than one year ago; quit within the past year; started to smoke < 1 year ago; have smoked for > 1 year), <u>drinking habit</u> (6–7 days/week; 4–5 days/week; 2–3 days/week; < 1 day/week; almost never), exercise habit, breakfast routine, time spent in one-way commuting and number of overtime hours worked/day. With regard to exercise, participants indicated the number of days each week on which they exercised for 30 minutes or more. Regarding breakfast, they indicated the number of days each week on which they had breakfast.

### Statistical analysis

Age- and sex-adjusted odds ratios (ORs) and multivariate adjusted ORs were analyzed using a multilevel logistic model with nesting by residential prefecture<u>^23^</u>. All analyses included the incidence rate of COVID-19 since declared pandemic status was an area-level variable. In the analyses of socioeconomic conditions, the model included sex, age, marital status, <u>equivalised</u> income <u>(divided into tertiles)</u>, job type, employment status, educational background, and level of comfort with financial condition. The model for analysis of health-related factors included age, sex, psychological distress, self-rated health, feeling of loneliness, having a friend who can provide support, and having a health condition which requires the employer’s support to allow work to occur. The model used to analyse lifestyle and work-related factors incorporated age, sex, <u>drinking habit</u>, smoking habit <u>(classified into three groups, “non-smoker”; “ex-smoker”; “current smoker”)</u>, exercise, breakfast routine, overtime hours worked/day and time required for one-way commuting.

A *P* value of less than .05 <u>(two-tailed)</u> was considered to indicate statistical significance. Stata Statistical Software version 16 (StataCorp LLC, TX, USA) was used for all analyses.

## Results

Basic participant characteristics are shown in Table 1. Among all 27,036 <u>participants</u>, approximately 19 percent of <u>participants</u> had experienced sickness presenteeism. On comparison, <u>participants</u> who had experienced sickness presenteeism were more likely than those who had not to have lower level of subjective health, greater psychological distress, greater loneliness, and greater need for employment consideration from their employer.

**Table 1.**
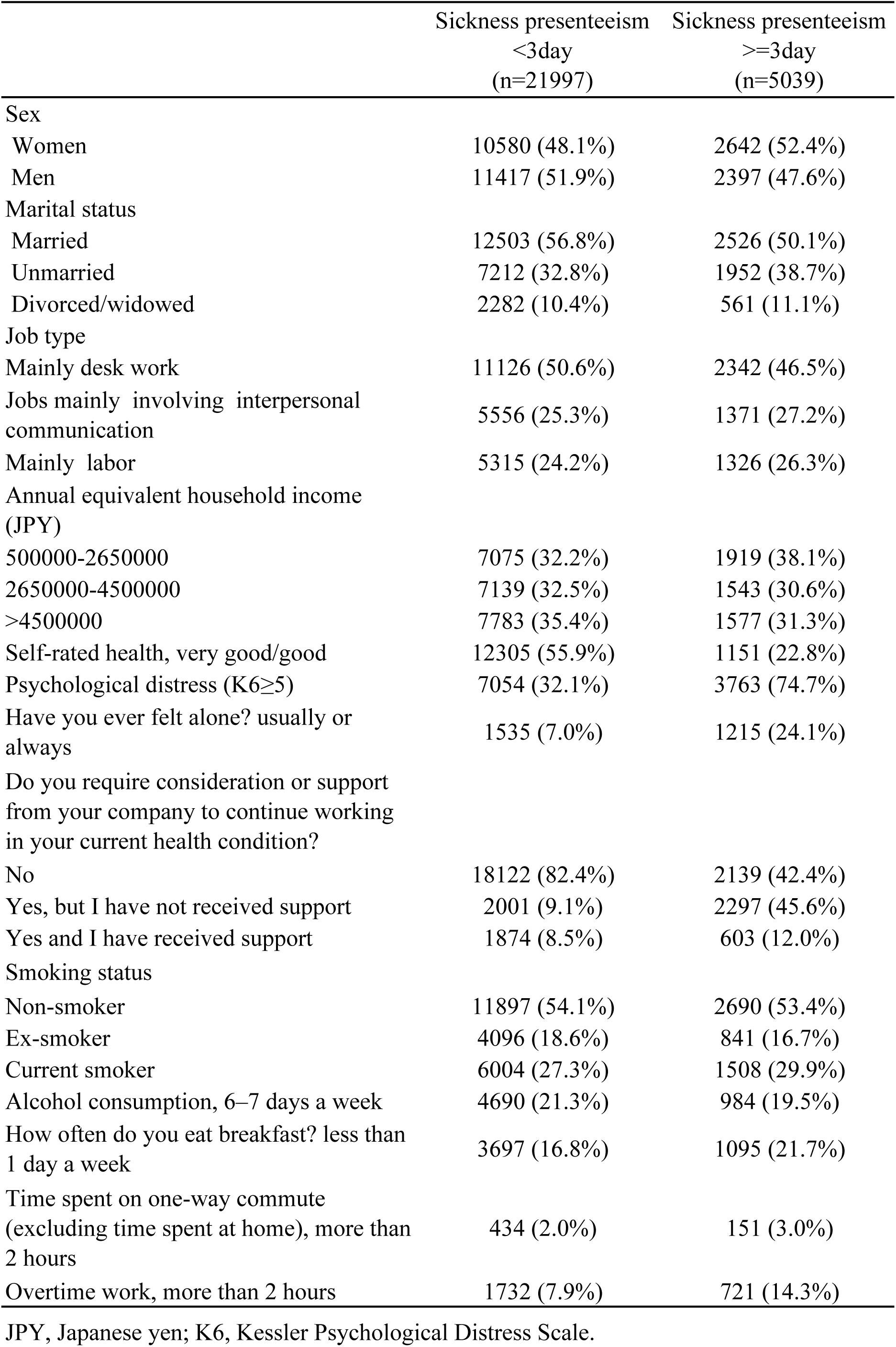
Basic characteristics of the study participants

|  | Sickness presenteeism<br><3day<br>(n=21997) | Sickness presenteeism<br>≥3day<br>(n=5039) |
| --- | --- | --- |
| Sex |  |  |
| Women | 10580 (48.1%) | 2642 (52.4%) |
| Men | 11417 (51.9%) | 2397 (47.6%) |
| Marital status |  |  |
| Married | 12503 (56.8%) | 2526 (50.1%) |
| Unmarried | 7212 (32.8%) | 1952 (38.7%) |
| Divorced/widowed | 2282 (10.4%) | 561 (11.1%) |
| Job type |  |  |
| Mainly desk work | 11126 (50.6%) | 2342 (46.5%) |
| Jobs mainly involving interpersonal communication | 5556 (25.3%) | 1371 (27.2%) |
| Mainly labor | 5315 (24.2%) | 1326 (26.3%) |
| Annual equivalent household income (JPY) |  |  |
| 500000-2650000 | 7075 (32.2%) | 1919 (38.1%) |
| 2650000-4500000 | 7139 (32.5%) | 1543 (30.6%) |
| >4500000 | 7783 (35.4%) | 1577 (31.3%) |
| Self-rated health, very good/good | 12305 (55.9%) | 1151 (22.8%) |
| Psychological distress (K6≥5) | 7054 (32.1%) | 3763 (74.7%) |
| Have you ever felt alone? usually or always | 1535 (7.0%) | 1215 (24.1%) |
| Do you require consideration or support from your company to continue working in your current health condition? |  |  |
| No | 18122 (82.4%) | 2139 (42.4%) |
| Yes, but I have not received support | 2001 (9.1%) | 2297 (45.6%) |
| Yes and I have received support | 1874 (8.5%) | 603 (12.0%) |
| Smoking status |  |  |
| Non-smoker | 11897 (54.1%) | 2690 (53.4%) |
| Ex-smoker | 4096 (18.6%) | 841 (16.7%) |
| Current smoker | 6004 (27.3%) | 1508 (29.9%) |
| Alcohol consumption, 6–7 days a week | 4690 (21.3%) | 984 (19.5%) |
| How often do you eat breakfast? less than 1 day a week | 3697 (16.8%) | 1095 (21.7%) |
| Time spent on one-way commute (excluding time spent at home), more than 2 hours | 434 (2.0%) | 151 (3.0%) |
| Overtime work, more than 2 hours | 1732 (7.9%) | 721 (14.3%) |

JPY, Japanese yen; K6, Kessler Psychological Distress Scale.

ORs of socioeconomic status associated with sickness presenteeism are shown in Table 2. On multivariate analysis, the <u>odds ratio (</u>OR) of sickness presenteeism associated with not being married versus being married was 1.15 <u>(95% confidence interval (95%CI): 1.04-1.28, p=0.009)</u>. Respective values were 1.11 <u>(95%CI: 1.03-1.20, p=0.009)</u> for manual labor versus desk work; 1.79 <u>(95%CI: 1.47-2.19, p<0.001)</u> and 2.29 <u>(95%CI: 1.79-2.92, p<0.001)</u> for lost employment when the COVID-19 pandemic started and continued unemployment compared with employment across the entire pandemic period; and 3.34 <u>(95%CI: 2.94-3.81, p<0.001)</u> for a feeling of financial instability versus stability.

**Table 2.**
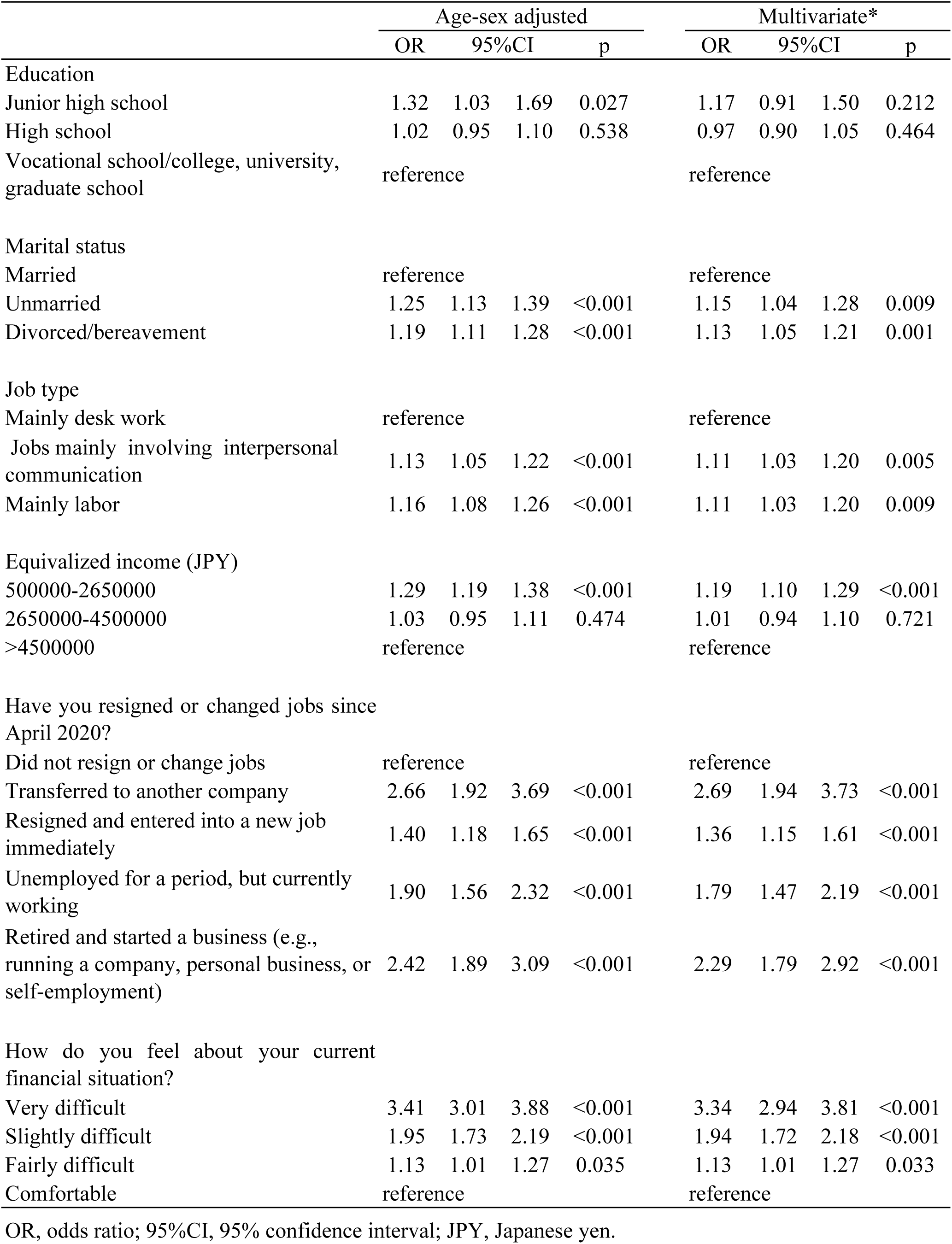
Association between socioeconomic status and sickness presenteeism

The ORs of health status and psychological distress associated with sickness presenteeism is shown in Table 3. On multivariate analysis, the OR of sickness presenteeism associated with poor self-rated health was 11.21 <u>(95%CI: 10.24-12.28, p<0.001)</u>; experience of psychological distress was 6.11 <u>(95%CI: 5.70-6.56, p<0.001)</u>; feeling always alone was 8.30 <u>(95%CI: 7.34-9.39, p<0.001)</u>; and the need for company consideration to allow work but not receiving it was 9.57 <u>(95%CI: 8.87-10.32, p<0.001)</u>.

**Table 3.**
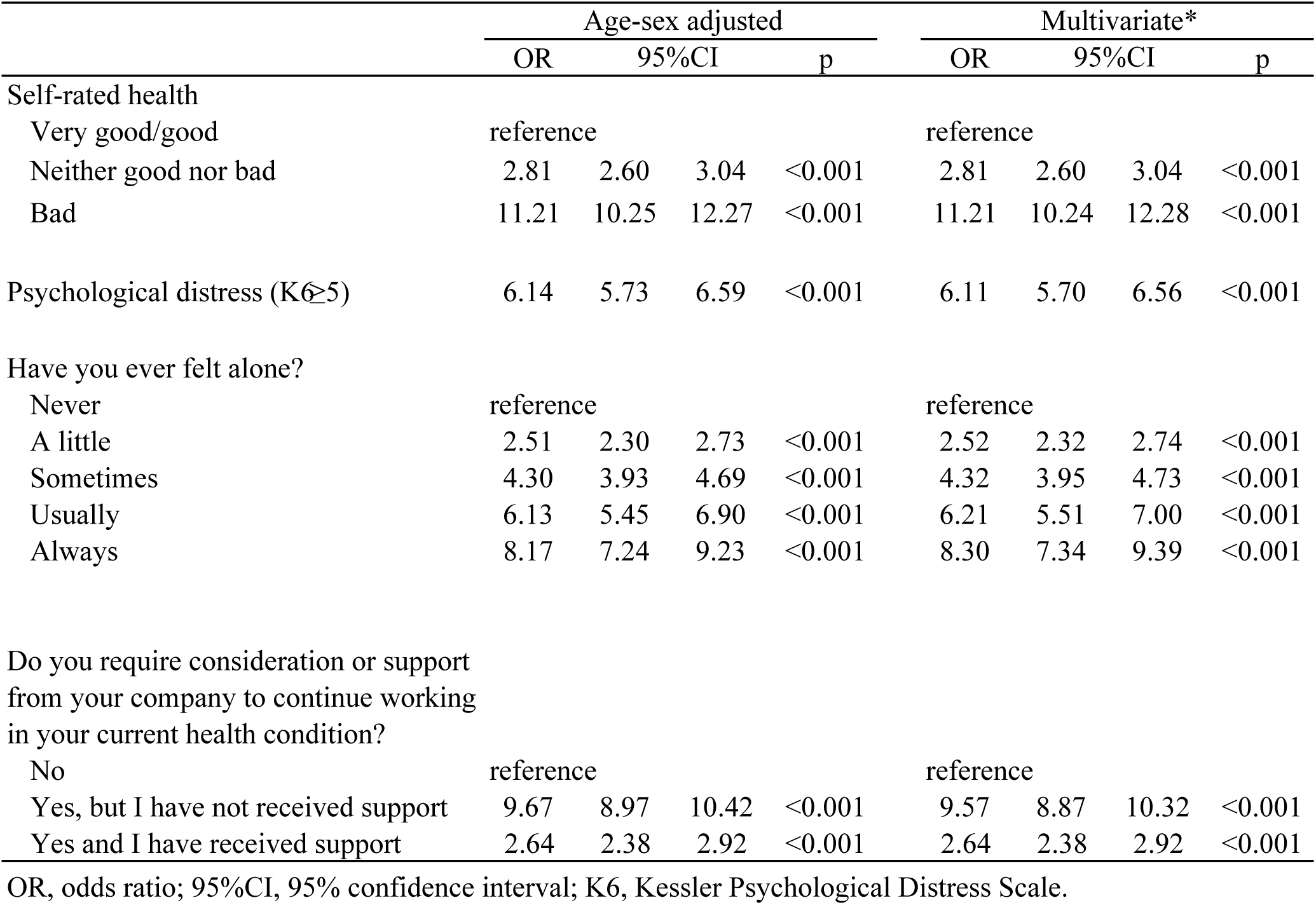
Association between health status, psychological distress and sickness presenteeism

|  | Age-sex adjusted |  |  |  | Multivariate* |  |  |  |
| --- | --- | --- | --- | --- | --- | --- | --- | --- |
|  | OR | 95%CI |  | p | OR | 95%CI |  | p |
| Self-rated health |  |  |  |  |  |  |  |  |
| Very good/good | reference |  |  |  | reference |  |  |  |
| Neither good nor bad | 2.81 | 2.60 | 3.04 | <0.001 | 2.81 | 2.60 | 3.04 | <0.001 |
| Bad | 11.21 | 10.25 | 12.27 | <0.001 | 11.21 | 10.24 | 12.28 | <0.001 |
| Psychological distress (K6≥5) | 6.14 | 5.73 | 6.59 | <0.001 | 6.11 | 5.70 | 6.56 | <0.001 |
| Have you ever felt alone? |  |  |  |  |  |  |  |  |
| Never | reference |  |  |  | reference |  |  |  |
| A little | 2.51 | 2.30 | 2.73 | <0.001 | 2.52 | 2.32 | 2.74 | <0.001 |
| Sometimes | 4.30 | 3.93 | 4.69 | <0.001 | 4.32 | 3.95 | 4.73 | <0.001 |
| Usually | 6.13 | 5.45 | 6.90 | <0.001 | 6.21 | 5.51 | 7.00 | <0.001 |
| Always | 8.17 | 7.24 | 9.23 | <0.001 | 8.30 | 7.34 | 9.39 | <0.001 |
| Do you require consideration or support from your company to continue working in your current health condition? |  |  |  |  |  |  |  |  |
| No | reference |  |  |  | reference |  |  |  |
| Yes, but I have not received support | 9.67 | 8.97 | 10.42 | <0.001 | 9.57 | 8.87 | 10.32 | <0.001 |
| Yes and I have received support | 2.64 | 2.38 | 2.92 | <0.001 | 2.64 | 2.38 | 2.92 | <0.001 |

OR, odds ratio; 95%CI, 95% confidence interval; K6, Kessler Psychological Distress Scale.

The ORs of lifestyle and occupational factors associated with sickness presenteeism is shown in Table 4. Smoking, breakfast routine, commute time, and hours worked overtime were associated with sickness presenteeism. In contrast, we saw no association between <u>drinking habit</u> (except less than 1 day a week) and sickness presenteeism. On multivariate analysis, current smokers were at higher likelihood of experiencing sickness presenteeism (OR=1.20, 95%CI: 1.12-1.29, p<0.001), as were those who rarely ate breakfast (OR=1.49, 95%CI: 1.38-1.62, p<0.001), spent longer than 2 hours in one-way commuting (OR=1.45, 95%CI: 1.18-1.77, p<0.001), or worked more than 2 hours overtime (OR=2.53, 95%CI: 2.53-2.80, p<0.001).

**Table 4.** Association between lifestyle, occupational factors and sickness presenteeism

|  | Age-sex adjusted |  |  |  | Multivariate* |  |  |  |
| --- | --- | --- | --- | --- | --- | --- | --- | --- |
|  | OR | 95%CI |  | p | OR | 95%CI |  | p |
| Smoking status |  |  |  |  |  |  |  |  |
| Non-smoker | reference |  |  |  | reference |  |  |  |
| Ex-smoker | 1.06 | 0.97 | 1.16 | 0.189 | 1.06 | 0.97 | 1.16 | 0.207 |
| Current smoker | 1.26 | 1.17 | 1.36 | <0.001 | 1.25 | 1.16 | 1.35 | <0.001 |
| Drinking habit |  |  |  |  |  |  |  |  |
| 6–7 days a week | 0.96 | 0.88 | 1.05 | 0.337 | 0.98 | 0.90 | 1.07 | 0.674 |
| 4–5 days a week | 0.91 | 0.80 | 1.03 | 0.142 | 0.94 | 0.83 | 1.06 | 0.301 |
| 2–3 days a week | 0.96 | 0.87 | 1.06 | 0.385 | 0.98 | 0.89 | 1.08 | 0.677 |
| Less than 1 day a week | 0.88 | 0.80 | 0.96 | 0.004 | 0.89 | 0.81 | 0.97 | 0.008 |
| Hardly ever | reference |  |  |  | reference |  |  |  |
| How often do you eat breakfast? |  |  |  |  |  |  |  |  |
| 6–7 days a week | reference |  |  |  | reference |  |  |  |
| 4–5 days a week | 1.43 | 1.29 | 1.59 | <0.001 | 1.41 | 1.27 | 1.57 | <0.001 |
| 2–3 days a week | 1.71 | 1.52 | 1.93 | <0.001 | 1.68 | 1.49 | 1.90 | <0.001 |
| Less than 1 day a week | 1.72 | 1.50 | 1.97 | <0.001 | 1.70 | 1.49 | 1.95 | <0.001 |
| Hardly ever | 1.54 | 1.42 | 1.66 | <0.001 | 1.49 | 1.38 | 1.62 | <0.001 |
| Time spent on one-way commute<br>(excluding time spent at home) |  |  |  |  |  |  |  |  |
| More than 2 hours | 1.35 | 1.11 | 1.66 | 0.003 | 1.45 | 1.18 | 1.77 | <0.001 |
| More than 1 hour | 1.06 | 0.95 | 1.19 | 0.272 | 1.13 | 1.01 | 1.27 | 0.029 |
| More than 30 minutes | 0.92 | 0.83 | 1.01 | 0.086 | 0.96 | 0.87 | 1.06 | 0.386 |
| Less than 30 minutes | 0.79 | 0.73 | 0.87 | <0.001 | 0.81 | 0.73 | 0.88 | <0.001 |
| Almost none | reference |  |  |  | reference |  |  |  |
| Overtime work |  |  |  |  |  |  |  |  |
| More than 2 hours | 2.39 | 2.16 | 2.64 | <0.001 | 2.53 | 2.28 | 2.80 | <0.001 |
| More than 1 hour | 1.67 | 1.53 | 1.83 | <0.001 | 1.75 | 1.60 | 1.91 | <0.001 |
| More than 30 minutes | 1.45 | 1.32 | 1.58 | <0.001 | 1.50 | 1.37 | 1.64 | <0.001 |
| Less than 30 minutes | 1.15 | 1.05 | 1.28 | 0.005 | 1.19 | 1.07 | 1.31 | 0.001 |
| Almost none | reference |  |  |  | reference |  |  |  |

OR, odds ratio; 95%CI, 95% confidence interval.

## Discussion

This study found that 19% of workers in Japan had experienced sickness presenteeism during the COVID-19 pandemic. <u>A survey of Japanese workers conducted before the COVID-19 pandemic found that 27.6% of workers had experienced sickness presenteeism due to mental health problems 2 to 5 times in the last 12 months^27^. It is not possible to make a simple comparison between this previous and the present study because the conditions of the two surveys are different. However, considering that the target period of this survey was only one month, it can be said that this result shows that COVID-19 is sufficiently associated with sickness presenteeism.</u> Risk of experiencing sickness presenteeism was higher in workers with a lower socioeconomic status and those in poor health. Further, risk was also higher in workers with an unfavorable lifestyle and working conditions.

Socioeconomic status is known to be an important factor in a worker’s experience of sickness presenteeism^14,<u>28</u>^. Further, emergencies such as disasters impair access to health care for people with a disadvantaged socioeconomic status<u>^29,30^</u>. As a result, opportunities for workers to experience sickness presenteeism can be expected to increase. The present study reveals that socioeconomically disadvantaged workers were more likely to experience sickness presenteeism during the COVID-19 pandemic. In particular, risk of sickness presenteeism was increased in subjects with low income, unemployment experience, and economic insecurity, a finding consistent with previous studies of the impact of COVID-19 on sickness presenteeism^21,22^. Economic deprivation is a direct reason for sickness presenteeism. Workers with unstable employment or a low income have little choice but to work hard for their livelihoods, even if they are not feeling well^14^. Conversely, workers with sickness presenteeism are more likely to exacerbate their illness^10^. As a result, sickness presenteeism will work in the direction of lower income and employment instability. Workers with unstable employment may not be entitled to sick leave or may be reluctant to take it<u>^31,32^</u>, rendering them prone to sickness presenteeism. Moreover, experience of sickness presenteeism has also been shown to exacerbate health status, resulting in turn in employment instability as well as economic disadvantage.

Our present results revealed that those with a poor health condition had a greater likelihood of experiencing sickness presenteeism: those whose self-rated health was poor, or with psychological distress, loneliness, and workplace support were at greater risk of experiencing sickness presenteeism. It is clear from the definition that sickness presenteeism is more likely in poor health^1^. Conversely, workers with sickness presenteeism are known to be more likely to exacerbate their illness^10^. In our present study, we found an association between greater psychological distress and a higher likelihood of sickness presenteeism. A number of studies have noted that the fear of infection when visiting a hospital is a major reason for treatment interruption<u>^33-35^</u>. Anxiety concerning infection may affect treatment discontinuation, resulting in continued working in poor physical condition. In addition, participants without company support were more likely to have sickness presenteeism. This result is consistent with reports that there is less sickness presenteeism with the support of workplace managers<u>^36^</u>. For those who tend to feel lonely, it is thought that work efficiency is likely to decline due to the infectious disease countermeasures (eg Stay Home and Remote Work) that have become widespread during the COVID-19 epidemic, and the resulting decline in workplace communication.

Our present results also revealed the <u>association</u> of non-preferable lifestyle and work-related factors with sickness presenteeism. Previous studies have shown a relationship between lifestyle and sickness presenteeism<u>^37-39^</u>. It is thought that stress has likely to have increased due to environmental changes caused by infectious disease countermeasures (eg, Stay Home and Remote Work) that have become widespread during the COVID-19 epidemic. Sickness presenteeism will increase due to a deterioration in lifestyle, such as the stress caused by countermeasures against COVID-19 and work at home, where the lack of restrictions on smoking leads to an increase in the number of cigarettes smoked<u>^40^</u>. <u>Long commute times put some physical burden on workers. Also, longer commute times reduce leisure and sleep time in daily life. Returning home late at night and having a supper also contribute to the worsening of lifestyle-related diseases. Therefore, long commute times can lead to sickness presenteeism^41^. Countermeasures against COVID-19 recommended in Japan such as staggered commuting and ventilation inside the train by opening windows require workers to go to work quite early in the morning and to travel in a train that is not adjusted to a comfortable temperature or humidity. This can also put a physical and mental burden on workers and increase sickness presenteeism. This is considered to be especially noticeable for workers who commute for long hours.</u>

Sickness presenteeism is a major occupational health issue and public health challenge. The COVID-19 pandemic has brought a particular focus to it. Our present study emphasizes that sickness presenteeism is associated with socioeconomic status. Regardless of causation, individuals with a lower socioeconomic or health status were at greater risk of sickness presenteeism. It is therefore critical to recognize not only the clinical importance of COVID-19, but in addition the deterioration in labor productivity and the impact on the lives and health of workers through the socioeconomic environment. <u>In a survey of Japanese workers, 62% of those with symptoms such as fever went to work within 7 days after symptom onset. Being hired by a company and not being able to work remotely were associated with going to work within 7 days of the onset of symptoms^42^</u>. It is important to foster the social consensus that workers should take the day off from work when they are feeling unwell. It is known that companies with a leave system that is easy to use when workers are not feeling well have a low incidence of infectious diseases^15^. An easy-to-use leave system is effective not only in reducing sickness presenteeism but also with regard to COVID-19 countermeasures.

There are several limitations in this study. First, the reasons for sickness presenteeism are not known. Many possible explanations may explain why workers experience sickness presenteeism, including financial hardship which prevents medical visits, unstable employment, or unavailability of a sick leave entitlement. Second, we did not investigate the types of illnesses and health problems experienced by people with sickness presenteeism. Third, we did not determined the chronological relationship between sickness presenteeism and socioeconomic status and lifestyle.

In this study, we found that around 19% of Japanese workers experienced sickness presenteeism during the period of rapidly spread of COVID-19 infection. Those who were socioeconomically disadvantaged, in poor health, or with an unfavorable lifestyle were more likely to report sickness presenteeism. Accordingly, the issue of sickness presenteeism has <u>become more prominent under the</u> COVID-19 <u>epidemic</u>. The increase in sickness presenteeism may not only worsen the health status of individuals, but also have long-term effects on society, such as reduced productivity and increased social security burden due to employment instability. These findings indicate the need for efforts to decrease sickness presenteeism in workers who need to recuperate at home.

## Data Availability

Data not available due to ethical restrictions.

## Acknowledgements

The current members of the CORoNaWork Project, in alphabetical order, are as follows: Dr. Yoshihisa Fujino (present chairperson of the study group), Dr. Akira Ogami, Dr. Arisa Harada, Dr. Ayako Hino, Dr. Hajime Ando, Dr. Hisashi Eguchi, Dr. Kazunori Ikegami, Dr. Kei Tokutsu, Dr. Keiji Muramatsu, Dr. Koji Mori, Dr. Kosuke Mafune, Dr. Kyoko Kitagawa, Dr. Masako Nagata, Dr. Mayumi Tsuji, Ms. Ning Liu, Dr. Rie Tanaka, Dr. Ryutaro Matsugaki, Dr. Seiichiro Tateishi, Dr. Shinya Matsuda, Dr. Tomohiro Ishimaru, and Dr. Tomohisa Nagata. All members are affiliated with the University of Occupational and Environmental Health, Japan.

## References

1. Aronsson G, Gustafsson K, Dallner M. Sick but yet at work. An empirical study of sickness presenteeism. J Epidemiol Community Health. 2000;54:502–509.

2. Eurofound. Health and well-being at work: a report based on the fifth european working conditions survey. Dublin: 2012.

3. Webster RK, Liu R, Karimullina K, et al. A systematic review of infectious illness Presenteeism: prevalence, reasons and risk factors. BMC Public Health. 2019 Jun 21;19:799.

4. Shan G, Wang S, Wang W, et al. Presenteeism in Nurses: Prevalence, Consequences, and Causes From the Perspectives of Nurses and Chief Nurses. Front Psychiatry. 2021 Jan 8;11:584040.

5. Park JW, Cho SS, Lee J, et al. Association between employment status and sickness presenteeism among Korean employees: a cross-sectional study. Ann Occup Environ Med. 2020 Jun 12;32:e17.

6. Oshio T, Tsutsumi A, Inoue A, et al. The reciprocal relationship between sickness presenteeism and psychological distress in response to job stressors: evidence from a three-wave cohort study. J Occup Health. 2017 Nov 25;59:552–561.

7. Taloyan M, Aronsson G, Leineweber C, et al. Sickness presenteeism predicts suboptimal self-rated health and sickness absence: A nationally representative study of the Swedish working population. PLoS One. 2012;7:e44721.

8. Kivimäki M, Head J, Ferrie JE, et al. Working while ill as a risk factor for serious coronary events: the Whitehall II study. Am J Public Health. 2005;95:98–102.

9. Conway PM, Hogh A, Rugulies R, et al. Is sickness presenteeism a risk factor for depression? A Danish 2-year follow-up study. J Occup Environ Med. 2014;56:595–603.

10. Hansen CD, Andersen JH. Sick at work-a risk factor for long-term sickness absence at a later date? J Epidemiol Community Health. 2009; 63:397–402.

11. Dellve L, Hadzibajramovic E, Ahlborg G Jr. Work attendance among healthcare workers: prevalence, incentives, and long-term consequences for health and performance. J Adv Nurs. 2011;67:1918–1929.

12. Rosvold EO, Bjertness E. Physicians who do not take sick leave: hazardous heroes? Scand J Public Health. 2001;29:71–75.

13. Johns G. Presenteeism in the workplace: A review and research agenda. Journal of Organizational Behavior 2010;31:519–542.

14. Aronsson G, Gustafsson K. Sickness presenteeism: Prevalence, attendance-pressure factors, and an outline of a model for research. J Occup Environ Med. 2005;47:958–966.

15. Böckerman P, Laukkanen E. What makes you work while you are sick? Evidence from a survey of workers. Eur J Public Health. 2010;20:43–46.

16. Allebeck P, Mastekaasa A. Chapter 5. Risk factors for sick leave - general studies. Scand J Public Health 2004;32:49–108.

17. Sverke M, Hellgren J, Näswall K. No security: a meta-analysis and review of job insecurity and its consequences. J Occup Health Psychol 2002;7:242–64.

18. Miraglia M, Johns G. Going to work ill: A meta-analysis of the correlates of presenteeism and a dual-path model. J Occup Health Psychol. 2016 Jul;21:261–283.

19. Bong CL, Brasher C, Chikumba E, et al. The COVID-19 Pandemic: Effects on Low-and Middle-Income Countries. Anesth Analg. 2020 Jul; 131: 86–92.

20. Das AK, Mishra DK, Gopalan SS. Reduced access to care among older American adults during CoVID-19 pandemic: results from a prospective cohort study. medRxiv. 2020: 2020.11.29.20240317.

21. Reuter M, Dragano N, Wahrendorf M. Working while sick in context of regional unemployment: a Europe-wide cross-sectional study. J Epidemiol Community Health. 2020 Nov 13:jech-2020-214888.

22. van Ballegooijen H, Goossens L, Bruin RH, et al. Concerns, quality of life, access to care and productivity of the general population during the first 8 weeks of the coronavirus lockdown in Belgium and the Netherlands. BMC Health Serv Res. 2021 Mar 12;21:227.

23. Fujino Y, Ishimaru T, Eguchi H, et al. Protocol for a nationwide Internet-based health survey in workers during the COVID-19 pandemic in 2020. medRxiv. Published online February 5, 2021:2021.02.02.21249309.

24. Kessler RC, Andrews G, Colpe LJ, et al. Short screening scales to monitor population prevalences and trends in non-specific psychological distress. Psychol Med. 2002;32:959–976.

25. Furukawa TA, Kawakami N, Saitoh M, et al. The performance of the Japanese version of the K6 and K10 in the World Mental Health Survey Japan. Int J Methods Psychiatr Res. 2008;17:152–158.

26. Courtin E, Knapp M. Social isolation, loneliness and health in old age: a scoping review. Health Soc Care Community. 2017 May;25:799–812.

27. Doki S, Sasahara S, Suzuki S, et al. Relationship between sickness presenteeism and awareness and presence or absence of systems for return to work among workers with mental health problems in Japan: an Internet-based cross-sectional study. J Occup Health. 2015;57:532–539.

28. Kim JY, Lee J, Muntaner C, et al. Who is working while sick? Nonstandard employment and its association with absenteeism and presenteeism in South Korea. Int Arch Occcup Environ Health. 2016;89:1095–1101.

29. Feinstein JS. The Relationship between Socioeconomic Status and Health: A Review of the Literature. M 323 ilbank Q. 1993;71:279–322.

30. Swain GR. How does economic and social disadvantage affect health. Focus. 2016;33:1–6.

31. Kinman G, Grant C. Presenteeism during the COVID-19 pandemic: risks and solutions. Occup Med. Published online November 18, 2020. doi:10.1093/occmed/kqaa193

32. Skagen K, Collins AM. The consequences of sickness presenteeism on health and wellbeing over time: A systematic review. Soc Sci Med. 2016;161:169–177.

33. Czeisler MÉ, Marynak K, Clarke KEN, et al. Delay or Avoidance of Medical Care Because of COVID-19-Related Concerns - United States, June 2020. MMWR Morb Mortal Wkly Rep. 2020;69:1250–1257.

34. Chudasama YV, Gillies CL, Zaccardi F, et al. Impact of COVID-19 on routine care for chronic diseases: A global survey of views from healthcare professionals. Diabetes Metab Syndr. 2020;14:965–967.

35. Erol MK. Treatment Delays and In-Hospital Outcomes In Acute Myocardial Infarction During The Covid-19 Pandemic: A Nationwide Study. The Anatolian Journal of Cardiology. Published online 2020. doi:10.14744/anatoljcardiol.2020.98607

36. Caverley N, Cunningham JB, MacGregor JN. Sickness presenteeism, sickness absenteeism, and health following restructuring in a public service organization. J Manag Studies. 2007;44:304–319.

37. Robroek SJ, van den Berg TI, Plat JF, et al. The role of obesity and lifestyle behaviours in a productive workforce. Occup Environ Med. 2011;68:134–139.

38. Kirkham HS, Clark BL, Bolas CA, et al. Which modifiable health risks are associated with changes in productivity costs? Popul Health Manag. 2015;18:30–38.

39. Merchant JA, Kelly KM, Burmeister LF, et al. Employment status matters: a statewide survey of quality-of-life, prevention behaviors, and absenteeism and presenteeism. J Occup Environ Med. 2014; 56:686–698.

40. Siegel A, Korbman M, Erblich J. Direct and Indirect Effects of Psychological Distress on Stress-Induced Smoking. J Stud Alcohol Drugs. 2017 Nov;78:930–937.

41. Min D, Lee SJ. Factors Associated With the Presenteeism of Single-Person Household Employees in Korea: The 5th Korean Working Conditions Survey (KWCS). Occup Environ Med. 2021 Sep 1;63:808–812.

42. Machida M, Nakamura I, Saito R, et al. The actual implementation status of self-isolation among Japanese workers during the COVID-19 outbreak. Trop Med Health. 2020 Aug 3;48:63. doi: 10.1186/s41182-020-00250-7.eCollection2020.

